# Factors influencing nurses’ susceptibility to online health misinformation: evidence from Greece

**DOI:** 10.1101/2025.11.19.25340604

**Authors:** Aglaia Katsiroumpa, Ioannis Moisoglou, Olympia Konstantakopoulou, Petros Galanis

## Abstract

**Background:** The ability to identify and counteract misinformation in healthcare is more critical than ever to safeguard public health and maintain confidence in evidence-based practices.

**Objective:** To identify predictors of online health misinformation in nurses. In particular, we examined the association between several demographic variables, trust in scientists and online health misinformation susceptibility.

**Methods:** A cross-sectional study was carried out in Greece, with data collected through an online survey conducted in October 2025. We used the Health-Related Online Misinformation Susceptibility Scale to measure online health misinformation susceptibility in our nurses. We used the Trust in Scientists Scale to measure levels of trust in scientists.

**Results:** We found that trust in scientists reduces online health misinformation susceptibility (adjusted coefficient beta = -3.290, 95% confidence interval [CI] = - 4.687 to -1.893, p<0.001). Moreover, we found that nurses with a MSc/PhD diploma had lower levels of misinformation susceptibility (adjusted coefficient beta = -2.470, 95% CI = -4.428 to -0.512, p=0.014). Additionally, interest in politics was associated with reduced misinformation susceptibility (adjusted coefficient beta = -0.794, 95% CI = -1.119 to -0.469, p<0.001).

**Conclusion:** Our findings showed that nurses who trusted scientists had also less online health misinformation susceptibility. Additionally, educational level and interest in politics affected online health misinformation susceptibility. Policy makers and healthcare organizations should reduce nurses’ misinformation susceptibility and improve their ability to detect fake news.

## Introduction

The digital era has revolutionized health information dissemination, with social media and online platforms becoming primary sources for both patients and healthcare professionals (Rolls & Massey, 2021). While these channels offer unprecedented access to medical knowledge, they also facilitate the rapid spread of misinformation, posing significant risks to public health and clinical practice (Wang et al., 2019). Health misinformation—defined as false or misleading health-related claims not supported by scientific evidence—has been linked to vaccine hesitancy, poor treatment adherence, and erosion of trust in healthcare systems (Kbaier et al., 2024).

Studies indicate that misinformation spreads faster and more broadly than accurate information on social media, driven by algorithms prioritizing engagement over accuracy and amplified by emotionally charged content. For instance, during the COVID-19 pandemic, misinformation about vaccines and treatments dominated online discourse, influencing patient decisions and creating additional burdens for healthcare professionals, including nurses, who often serve as trusted sources of health guidance (Lee et al., 2022; Pierri et al., 2022; Rodrigues et al., 2023).

Nurses, often regarded as trusted health professionals, play a critical role in patient education and evidence-based care. However, research indicates that nurses themselves can be susceptible to misinformation due to information overload, low digital health literacy, and cognitive biases. Algorithm-driven echo chambers and confirmation bias reinforce false beliefs, making it harder for nurses to discern credible sources (Deori et al., 2025; Sharman, 2023). The proliferation of online health misinformation poses significant challenges for healthcare professionals, including nurses, who play a pivotal role in patient education and evidence-based practice.

The COVID-19 pandemic was accompanied by an unprecedented surge in health misinformation, leading the World Health Organization (WHO) to describe the phenomenon as an “infodemic”—a situation where excessive amounts of accurate and inaccurate information circulate simultaneously, making it difficult for individuals to identify trustworthy sources (The Lancet Infectious Diseases, 2020). In particular, social media platforms played a central role in accelerating the spread of misinformation. Algorithmic systems designed to maximize engagement often prioritized sensational or emotionally charged content over verified information, enabling false narratives to reach global audiences rapidly (Cinelli, Quattrociocchi, et al., 2020). Automated bots and influencers further amplified misleading claims, creating echo chambers that reinforced misinformation (Islam et al., 2020).

Thus, the COVID-19 pandemic illustrates the profound influence of the contemporary information ecosystem. The rapid dissemination of content significantly shapes individual behaviors and can undermine the effectiveness of government interventions. Social media platforms grant users direct access to vast amounts of information, often amplifying rumors and questionable claims. Algorithms, designed to reflect user preferences and attitudes, facilitate the promotion and circulation of content, thereby accelerating information spread (Kulshrestha et al., 2017). This departure from traditional news paradigms has far-reaching implications for the formation of social perceptions and narrative framing, influencing policy-making, political discourse, and the trajectory of public debate—particularly on contentious issues (Del Vicario, Bessi, et al., 2016; Schmidt et al., 2017; Starnini et al., 2016). Online users frequently seek information that aligns with their existing beliefs, disregard opposing viewpoints, and form polarized communities around shared narratives (Baronchelli, 2018; Cinelli, Brugnoli, et al., 2020). Moreover, in highly polarized environments, misinformation can proliferate with ease (Bail et al., 2018; Del Vicario, Vivaldo, et al., 2016).

In this context, online health misinformation is a significant concern for nurses because it impacts directly on their core roles of healthcare providers and patient educators. Nurses occupy a unique position as frontline educators and advocates for evidence-based care (Makic, 2025). Central to this role is assisting individuals to make more informed health-related decisions by evaluating and improving their health literacy (McCaskill et al., 2024). Therefore, nurses’ digital health literacy—the ability to seek, understand, and evaluate health information online—is a critical protective factor. Integrating digital literacy into lifelong learning for healthcare professionals to counter misinformation effectively is crucial to combat online health misinformation. Interventions such as educational programs, psychological inoculation, and source credibility labeling have shown promise, though evidence on their long-term effectiveness remains mixed (Grover et al., 2025; Wamala Andersson & Gonzalez, 2025).

However, the existing literature on measuring online health misinformation among nurses remains notably scarce. Although health misinformation is increasingly recognized as a critical challenge within healthcare education, there is a clear lack of empirical studies that quantify its prevalence or evaluate susceptibility in this specific population. Therefore, the aim of our study was to examine potential predictors of online health misinformation in nurses. In particular, we examined the association between several demographic variables, trust in scientists and online health misinformation susceptibility.

## Methods

### Study design

A cross-sectional study was conducted in Greece, with data collection taking place via an online survey in October 2025. The study questionnaire was developed using Google Forms and disseminated through Facebook and Instagram groups targeting nurses. Additionally, we distributed the survey by sending direct messages to nurses on LinkedIn. This approach yielded a convenience sample. Eligible participants had to meet the following criteria: (1) be employed as clinical nurses in healthcare settings, (2) spend at least 30 minutes per day on the web or social media, and (3) provide informed consent to participate. The study adhered to the Strengthening the Reporting of Observational Studies in Epidemiology (STROBE) guidelines (Von Elm et al., 2008).

We used G*Power v.3.1.9.2 to calculate our sample size. We included nine predictors in our multivariable models. Thus, considering an anticipated effect size of 0.04 between each independent variable and outcome (online health misinformation susceptibility), a statistical power of 95%, and a margin of error of 5%, the sample size was estimated to be 327 nurses.

### Measurements

We measured several demographic characteristics of nurses: gender (males or females), age (continuous variable), MSc/PhD diploma (no or yes), financial status (self-assessment scale from 0 [very poor financial status] to 10 [excellent financial status]), level of trust in websites to report the news with integrity (self-assessment scale from 0 [not at all] to 10 [completely]), level of interest in politics (self-assessment scale from 0 [not at all] to 10 [completely]), and daily time in web/social media (continuous variable in hours).

We used the Health-Related Online Misinformation Susceptibility Scale (HR-OMISS) (Katsiroumpa, Konstantakopoulou, et al., 2025) to measure online health misinformation susceptibility in our nurses. The HR-OMISS is an adapted version of the Online Misinformation Susceptibility Scale (OMISS) (Katsiroumpa, Moisoglou, Mangoulia, et al., 2025) that measures online misinformation susceptibility in general, while the HR-OMISS measures specifically online health misinformation susceptibility. The HR-OMISS includes nine items such as “When you see a health-related post or story that interests you on social media or websites, how often do you check the website domain and URL?” and “When you see a health-related post or story that interests you on social media or websites, how often do you check the publication date of the post?”. Answers are on a five-point Likert scale; never (5), rarely (4), sometimes (3), very often (2), always (1). Total score ranges from 9 to 45. Higher scores indicate higher misinformation susceptibility. Developers of the OMISS suggest a cut-off point (≥23) (Katsiroumpa, Moisoglou, Gallos, et al., 2025) to distinguish individuals that show high levels of misinformation susceptibility from those who show normal levels of misinformation susceptibility. We used the valid Greek version of the HR-OMISS (Katsiroumpa, Konstantakopoulou, et al., 2025). In our study, the Cronbach’s alpha for the HR-OMISS was 0.929.

To assess nurses’ trust in scientists, we employed the Trust in Scientists Scale (TISS) (Cologna et al., 2025), which consists of 12 items designed to measure four key dimensions: integrity, competence, benevolence and openness. Given the high correlation among these dimensions, we selected integrity as a representative factor for use in our study. The integrity scale includes three items: How honest or dishonest are most scientists?, How ethical or unethical are most scientists?, and How sincere or insincere are most scientists?. Responses are recorded on a five-point Likert scale, ranging from 1 (very dishonest/unethical/insincere) to 5 (very honest/ethical/sincere). The integrity score was computed as the mean of the three item scores, resulting in a total score ranging from 1 to 5. Higher scores reflect greater trust in scientists. We used the valid Greek version of the Trust in Scientists Scale (Cologna et al., 2025). In our study, Cronbach’s alpha for the integrity scale was 0.913.

### Ethical issues

We conducted our study in accordance with the Declaration of Helsinki (“World Medical Association Declaration of Helsinki,” 2013). Moreover, the Ethics Committee of the Faculty of Nursing, National and Kapodistrian University of Athens approved our study protocol (approval No. 75, July 13, 2025). We collected our data on an anonymous and voluntary basis. We informed nurses about the aim and the design of our study, and they gave their informed consent.

### Statistical analysis

We present categorical variables as numbers and percentages. Also, we use mean, standard deviation (SD), median, minimum value, and maximum value to present continuous variables. We used the Kolmogorov-Smirnov test and Q-Q plots to examine the distribution of continuous variables. We found that continuous variables followed normal distribution. Demographic variables and trust in scientists were the independent variables, while online health misinformation susceptibility was the dependent variable. Since our dependent variable was continuous variable that followed normal distribution we employed linear regression analysis. First, we performed simple linear regression analysis, and then we constructed a final multivariable model to estimate the independent effect of each independent variable on outcomes. We present unadjusted and adjusted coefficients beta, 95% confidence intervals (CI), and p-values. We calculated Pearson’s correlation coefficient to examine correlations between the study scales since scores on these scales followed normal distribution. P-values less than 0.05 were considered statistically significant. We used the IBM SPSS 28.0 (IBM Corp. Released 2021. IBM SPSS Statistics for Windows, Version 28.0. Armonk, NY: IBM Corp) for the analysis.

## Results

### Demographic characteristics

Table 1 shows demographic characteristics of the study sample. Our study sample included 373 nurses. Among them, 81.5% were females, and 72.7% held a MSc/PhD diploma. Mean age was 41.98 years (SD; 10.38) with a median of 43 years, a minimum age of 24 years and a maximum age of 67 years. Mean score of financial status was 5.96 (SD; 1.46) while median score was 6 (range; 0 to 10). Mean trust score in websites was 3.69 (SD; 1.97) with a median of 4 and a range between 0 and 9. Mean score of interest in politics was 5.23 (SD; 2.75), while median score was 6 (range; 0 to 10). Mean daily time in web/social media was 2.94 hours (SD; 2.13) with a median of 2.5 hours, a minimum of 30 minutes and a maximum of 8 hours.

**Table 1.** Demographic characteristics of the study sample (N=373).

| Characteristics | N | % |
| --- | --- | --- |
| Gender |  |  |
| Males | 69 | 18.5 |
| Females | 304 | 81.5 |
| Age<sup>a</sup> | 41.98 | 10.38 |
| MSc/PhD diploma |  |  |
| No | 102 | 27.3 |
| Yes | 271 | 72.7 |
| Financial status<sup>a</sup> | 5.96 | 1.46 |
| Trust in websites<sup>a</sup> | 3.69 | 1.97 |
| Interest in politics<sup>a</sup> | 5.23 | 2.75 |
| Daily time in web/social media (hours)<sup>a</sup> | 2.94 | 2.13 |

<sup>a</sup> mean, standard deviation

### Study scales

Table 2 presents descriptive statistics for the study scales. Mean score on HR-OMISS was 26.29 (SD; 8.89) with a median of 26. Also, 63.0% (n=235) of our nurses experienced high levels of online health misinformation susceptibility. Mean score on TISS was 3.25 (SD; 0.66).

**Table 2.**
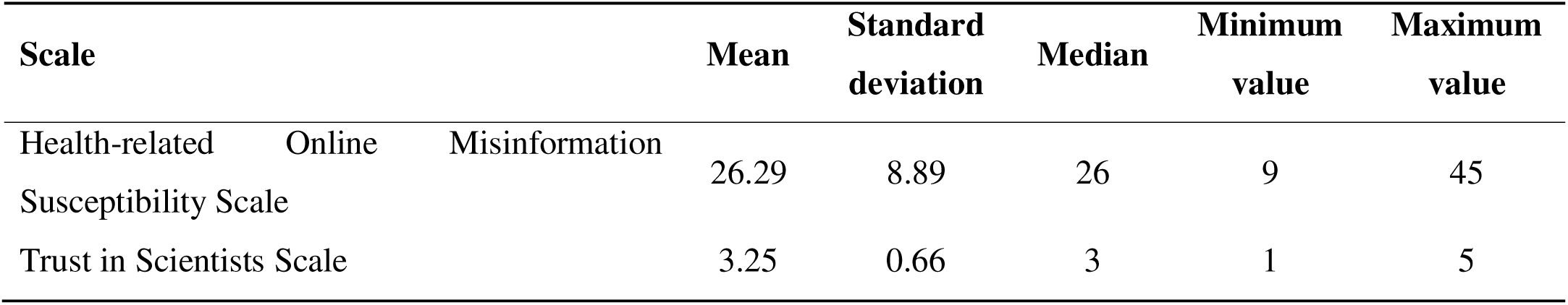
Descriptive statistics for the study scales (N=373).

We found a negative correlation between online misinformation susceptibility and trust in scientists (r = -0.320, p-value < 0.001).

### Dependent variable: online health misinformation susceptibility

Table 3 shows linear regression models with HR-OMISS as the dependent variable. We found that trust in scientists reduces online health misinformation susceptibility (adjusted coefficient beta = -3.290, 95% CI = -4.687 to -1.893, p<0.001). In other words, nurses who trusted scientists had also less online health misinformation susceptibility. Moreover, we found that nurses with a MSc/PhD diploma had lower levels of misinformation susceptibility (adjusted coefficient beta = -2.470, 95% CI = - 4.428 to -0.512, p=0.014). Additionally, interest in politics was associated with reduced misinformation susceptibility (adjusted coefficient beta = -0.794, 95% CI = - 1.119 to -0.469, p<0.001).

**Table 3.**
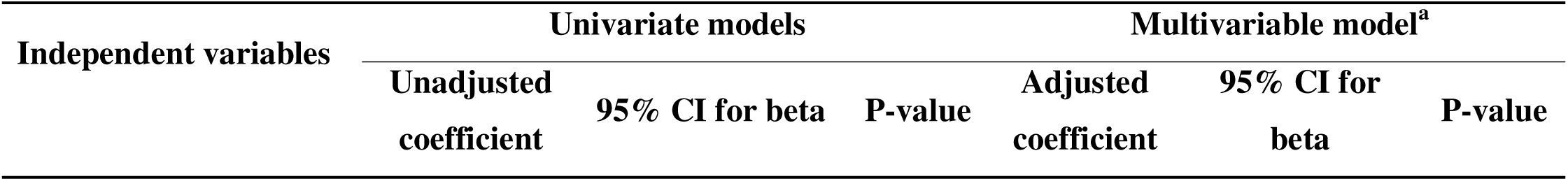
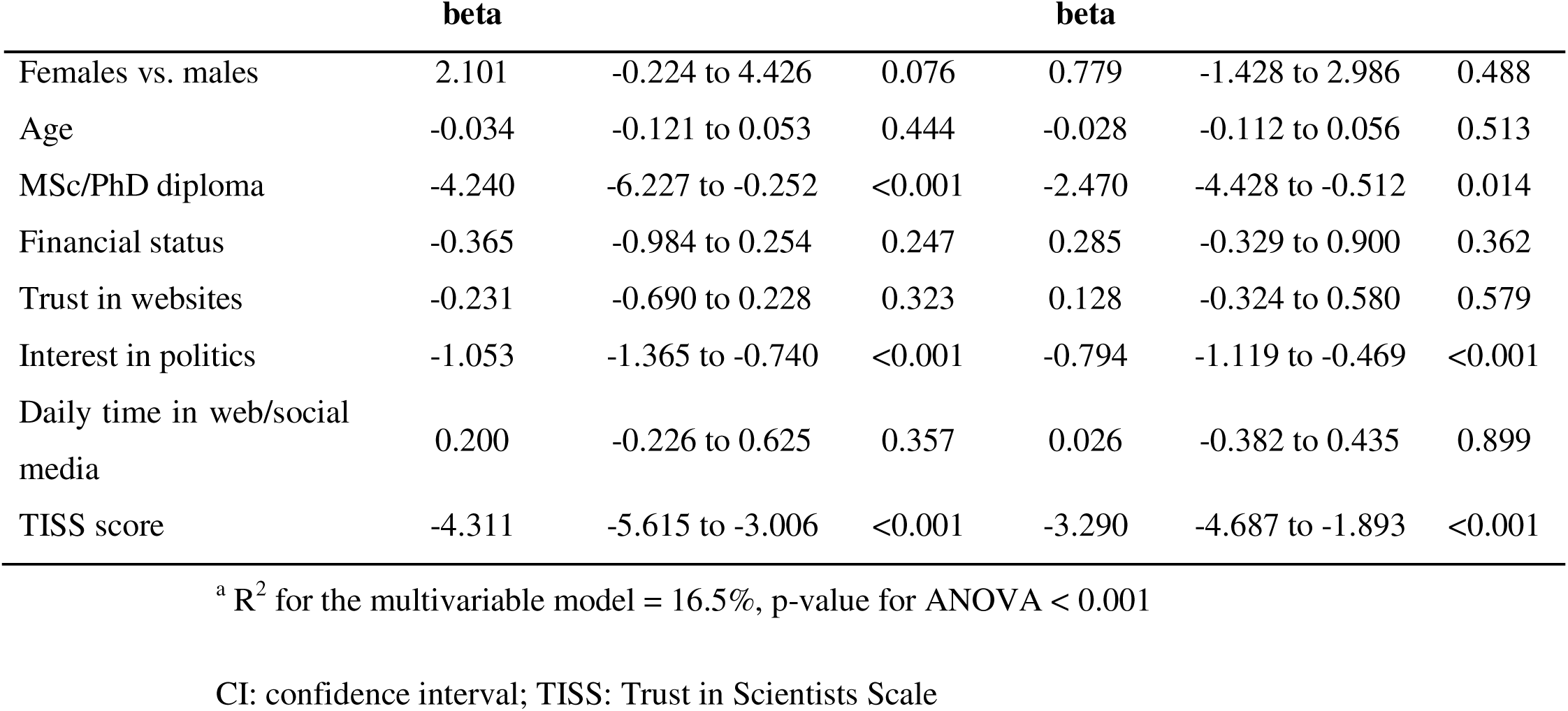
Linear regression models with Health-related Online Misinformation Susceptibility Scale as the dependent variable (N=373).

| Independent variables | Univariate models |  |  | Multivariable model<sup>a</sup> |  |  |
| --- | --- | --- | --- | --- | --- | --- |
|  | Unadjusted coefficient | 95% CI for beta | P-value | Adjusted coefficient | 95% CI for beta | P-value |

|  | **beta** |  |  | **beta** |  |  |
| --- | --- | --- | --- | --- | --- | --- |
| Females vs. males | 2.101 | -0.224 to 4.426 | 0.076 | 0.779 | -1.428 to 2.986 | 0.488 |
| Age | -0.034 | -0.121 to 0.053 | 0.444 | -0.028 | -0.112 to 0.056 | 0.513 |
| MSc/PhD diploma | -4.240 | -6.227 to -0.252 | <0.001 | -2.470 | -4.428 to -0.512 | 0.014 |
| Financial status | -0.365 | -0.984 to 0.254 | 0.247 | 0.285 | -0.329 to 0.900 | 0.362 |
| Trust in websites | -0.231 | -0.690 to 0.228 | 0.323 | 0.128 | -0.324 to 0.580 | 0.579 |
| Interest in politics | -1.053 | -1.365 to -0.740 | <0.001 | -0.794 | -1.119 to -0.469 | <0.001 |
| Daily time in web/social media | 0.200 | -0.226 to 0.625 | 0.357 | 0.026 | -0.382 to 0.435 | 0.899 |
| TISS score | -4.311 | -5.615 to -3.006 | <0.001 | -3.290 | -4.687 to -1.893 | <0.001 |

<sup>a</sup> R<sup>2</sup> for the multivariable model = 16.5%, p-value for ANOVA < 0.001
CI: confidence interval; TISS: Trust in Scientists Scale

## Discussion

Identifying the factors that influence nurses’ susceptibility to online health misinformation is of paramount importance, given their critical role as frontline healthcare professionals responsible for promoting health, delivering evidence-based care, and guiding individuals toward informed health-related decisions. As trusted sources of information, nurses significantly shape patient understanding and adherence to accurate health practices; therefore, recognizing and addressing the determinants of misinformation vulnerability is essential to safeguard public health and maintain confidence in healthcare systems.

In this context, the present study was designed to evaluate the level of online health misinformation susceptibility among nurses and to explore potential predictors associated with its occurrence. Multivariable analysis showed that trust in scientists, educational level and interest in politics reduce online health misinformation susceptibility in nurses.

The finding that 63.0% of nurses in our sample exhibited high levels of susceptibility to online health misinformation highlights a critical challenge for nursing practice and healthcare communication. This substantial proportion suggests that misinformation is not merely a public issue but also affects healthcare professionals who are expected to serve as reliable sources of evidence-based information (Makic, 2025). Nurses are frontline educators and trusted sources of health information; therefore, their vulnerability to misinformation has implications not only for their own decision-making but also for patient education and public health outcomes. Several factors may explain this vulnerability. First, the increasing reliance on digital platforms for professional updates and patient education exposes nurses to vast amounts of unverified content, often amplified by social media algorithms that prioritize engagement over accuracy. Second, gaps in digital health literacy and limited training in critical appraisal skills may hinder nurses’ ability to differentiate credible sources from misleading information. Cognitive biases, such as confirmation bias, and the emotional appeal of misinformation further exacerbate susceptibility.

Our findings indicated that increased trust in science significantly reduces nurses’ susceptibility to online health misinformation. This result aligns with prior research emphasizing the role of institutional trust as a protective factor against misinformation (Bhattacharya & Singh, 2025; Scherer & Pennycook, 2020). Trust in science functions as a cognitive anchor, guiding individuals toward evidence-based sources and away from unverified claims. Nurses who exhibit high trust in scientific processes are more likely to engage in critical appraisal, seek corroboration from peer-reviewed literature, and reject anecdotal or pseudoscientific narratives (Wamala Andersson & Gonzalez, 2025). From a psychological perspective, trust in science enhances epistemic vigilance, reducing reliance on heuristics and mitigating biases such as the illusory truth effect and motivated reasoning, which often lead individuals to accept repeated or belief-congruent misinformation (Bhattacharya & Singh, 2025). Conversely, low trust in science creates an environment where misinformation thrives, as individuals may perceive scientific recommendations as biased or politically motivated, making alternative narratives more appealing (Deori et al., 2025).

Our study found that higher educational attainment among nurses significantly reduces susceptibility to online health misinformation. This finding is consistent with prior research indicating that education enhances critical thinking skills, scientific reasoning, and the ability to evaluate evidence-based sources (Alotaibi et al., 2025; Scherer & Pennycook, 2020). Education provides nurses with a stronger foundation in health sciences, enabling them to recognize inaccuracies and apply professional judgment when encountering questionable information online (Sharman, 2023). From a cognitive perspective, higher education fosters analytical processing rather than heuristic shortcuts, reducing reliance on superficial cues such as source familiarity or emotional appeal. It also mitigates the illusory truth effect, as educated individuals are more likely to question repeated claims and seek corroboration from credible sources (Bhattacharya & Singh, 2025). Furthermore, advanced education often includes exposure to research methodology and evidence-based practice, which strengthens epistemic vigilance and skepticism toward unverified content.

This study revealed that greater interest in politics is associated with lower susceptibility to health misinformation among nurses. It seems to be that individuals who actively follow political issues tend to consume diverse sources of information and develop habits of evaluating competing claims (Scherer & Pennycook, 2020). These behaviors foster analytical thinking and skepticism toward unverified content, which can extend beyond political topics to health-related information (Bhattacharya & Singh, 2025). Political engagement often correlates with higher media literacy, as politically interested individuals are more accustomed to identifying bias, verifying sources, and questioning narratives (Deori et al., 2025). This vigilance can reduce reliance on heuristics and mitigate cognitive biases such as the illusory truth effect, which makes repeated misinformation seem credible. Furthermore, political interest may reflect a broader orientation toward civic responsibility and trust in institutional processes, including science and evidence-based policy, which reinforces resistance to misinformation (Wamala Andersson & Gonzalez, 2025).

Several limitations should be considered when interpreting this study’s findings. First, the sample consisted solely of nurses, which may restrict the applicability of results to other healthcare fields from diverse cultural and educational backgrounds. Future studies using random, representative samples across different countries and healthcare professionals would provide more robust insights. Second, although a validated scale was employed to assess misinformation susceptibility, the study did not account for differences in platform algorithms or content exposure, which could shape individual experiences. Third, the use of a cross-sectional design limits the ability to draw causal conclusions between demographic characteristics, trust in scientists, and vulnerability to online health misinformation. Longitudinal research is needed to clarify temporal patterns and causality. Fourth, reliance on self-reported data introduces potential social desirability bias and inaccuracies in reporting behaviors such as time spent on social media. Lastly, the focus of this study was on demographic factors, leaving out other relevant variables such as cognitive styles, digital literacy, and psychological traits. Future research should incorporate these dimensions for a more comprehensive understanding of misinformation vulnerability.

## Conclusions

This study was undertaken to assess the level of health misinformation susceptibility among nurses and to identify potential predictors associated with its occurrence. By systematically measuring misinformation levels and examining influencing factors, we aim to provide evidence that can inform targeted interventions, enhance digital health literacy, and strengthen nurses’ capacity to deliver accurate, evidence-based information in clinical and public health contexts.

Our findings carry significant implications since nurses who internalize inaccurate information may inadvertently transmit it to patients, undermining public trust and compromising health outcomes. Therefore, targeted interventions—such as structured digital literacy programs, integration of misinformation detection strategies into nursing curricula, and institutional policies promoting evidence-based resources—are urgently needed to strengthen nurses’ resilience against misinformation and safeguard the integrity of healthcare delivery.

In particular, digital health literacy training is a cornerstone intervention, emphasizing skills in evaluating source credibility, identifying misinformation patterns, and using fact-checking tools and evidence-based databases. Integrating these programs into nursing curricula and continuing education can ensure that nurses are equipped to navigate complex online environments (McCaskill et al., 2024; Wilandika et al., 2023).

In addition, critical thinking and cognitive bias awareness programs can help nurses recognize psychological factors—such as confirmation bias—that contribute to misinformation acceptance. Simulation-based learning and case-based exercises using real-world misinformation scenarios have been shown to improve decision-making under uncertainty (Anselmann et al., 2024; Muscat et al., 2021).

Institutional support is equally important. Healthcare organizations should provide verified repositories of health information and establish clear guidelines for professional use of social media to prevent inadvertent dissemination of false content. Collaboration with fact-checking organizations and public health authorities can further enhance access to accurate information and promote trust in official sources.

These interventions collectively aim to reduce susceptibility to misinformation, strengthen professional competence, and maintain public confidence in healthcare systems.

## Data Availability

All data produced are available online at figshare at https://doi.org/10.6084/m9.figshare.30661352

https://doi.org/10.6084/m9.figshare.30661352

## Notes

### Competing Interest Statement

The authors have declared no competing interest.

### Funding Statement

This study did not receive any funding

### Author Declarations

the Ethics Committee of the Faculty of Nursing, National and Kapodistrian University of Athens approved our study protocol (approval No. 75, July 13, 2025)

